# Prediction of Subsolid Pulmonary Nodule Evolution from Baseline CT Using Temporal Imaging Models

**DOI:** 10.64898/2026.08.12.26360292

**Authors:** Masha Bondarenko, Kang Qi, Ali Nowroozi, Justin Kim, Brian Kunzang, Aaron Lee, Jonathan Liu, Nicole Tran, Shiny Weng, Maya Vella, Gunvant Chaudhari, Tician Schnizler, Arun Innanje, Terrence Chen, Jae Ho Sohn

**Affiliations:** University of California, San Francisco (UCSF), Department of Radiology and Biomedical Imaging, San Francisco, USA; Peking University First Hospital, Beijing, China; University of California, Berkeley, USA; Institute for Diagnostic and Interventional Radiology, Cantonal Hospital Aarau (KSA), Aarau, Switzerland; United Imaging Intelligence (UII), Boston, USA

**Author notes:** **Corresponding Author Contact:** Jae Ho Sohn, MD, MS, **Email:**, **Address:** University of California, San Francisco Department of Radiology and Biomedical Imaging 185 Berry Street, Suite 350, Lobby 6, San Francisco, CA 94107.

**Keywords:** Subsolid pulmonary nodules, Computed tomography, Longitudinal imaging, Growth prediction, Deep learning, Generative modeling, Pulmonary adenocarcinoma

## Abstract

**Background:** Prediction of subsolid pulmonary nodule (SSN) progression from baseline CT may improve risk stratification and surveillance planning, but prior approaches have largely relied on fixed follow-up intervals.

**Methods:** This retrospective single-center study evaluated interval-aware temporal imaging models for predicting future SSN growth and morphology across heterogeneous surveillance durations. A total of 24,946 longitudinal scan pairings derived from 2,543 clinician-reviewed SSNs in 426 patients were analyzed. A discriminative deep learning model predicted interval growth from baseline CT, segmentation masks, and interscan interval information, while a temporally conditioned generative model predicted future lesion morphology.

**Results:** The discriminative model achieved an area under the receiver operating characteristic curve of 0.772 (95% confidence interval: 0.704–0.818), with sensitivity of 80.2% and specificity of 58.7% on the test cohort. The generative model predicted future lesion morphology with a Dice similarity coefficient of 0.706 ± 0.186. Prediction performance decreased with increasing follow-up duration, although both models generalized across intervals ranging from months to years.

**Conclusion:** Interval-aware temporal imaging models enable the prediction of future SSN growth and morphology from baseline CT while accounting for variable surveillance intervals. These findings suggest a framework for time-aware, personalized risk assessment that may support individualized surveillance strategies and future AI-assisted management of pulmonary adenocarcinoma spectrum lesions.

## 1. Introduction

Subsolid nodules (SSNs), a subtype of pulmonary nodules, are a critical focus in radiology due to their association with early-stage lung adenocarcinomas and their potential for malignancy (1–3). These nodules, characterized by slow growth and mostly indolent behavior, present challenges in clinical management, requiring a balance between overtreatment of indolent lesions and delayed detection of malignant ones (1–9).

Prediction of future SSN progression from baseline imaging could improve management by identifying lesions more likely to demonstrate interval growth while reducing surveillance burden for stable nodules. Prior studies have demonstrated that baseline CT features, radiomics, and deep learning methods may predict future SSN growth at fixed follow-up timepoints, malignancy risk, or volume doubling time (10–13). However, such approaches may not fully reflect clinical practice, where surveillance intervals vary substantially across patients and lesions. Consequently, most prior SSN prediction studies have evaluated growth at fixed follow-up horizons rather than explicitly modeling lesion progression across heterogeneous surveillance intervals encountered in clinical practice.

Temporal modeling of SSN evolution presents additional challenges as lesion progression occurs non-linearly and involves both volumetric and spatial morphologic change. Most conventional prediction approaches summarize progression using categorical (e.g., growth vs no growth) or scalar (e.g., final size) outcomes, which may incompletely characterize the evolving geometry of lesions observed on serial CT examinations (10,11). Generative imaging approaches have increasingly been explored in radiology for image synthesis, reconstruction, and predictive modeling because they enable modeling of spatial image structure as well as categorical outcomes (14–22). Although prior studies have explored generative approaches for pulmonary nodule characterization and future imaging prediction, these methods have largely focused on malignancy classification or prediction at predefined temporal intervals (11,14). To our knowledge, no prior study has evaluated interval-aware generative prediction of future SSN morphology across heterogeneous longitudinal surveillance intervals.

Together, these limitations suggest that complementary approaches capable of modeling both interval growth rate and future lesion morphology may provide a more comprehensive representation of SSN progression. Accordingly, this study evaluated two temporal interval-aware imaging models for forecasting SSN progression from baseline CT across heterogeneous follow-up durations: a classification model for interval growth prediction and a generative segmentation model for estimation of future lesion morphology.

## 2. Materials and Methods

### Study Population

This retrospective single-institution study was approved by the institutional review board with waiver of written informed consent and was compliant with the Health Insurance Portability and Accountability Act. Consecutive patients undergoing chest CT between 2015 and 2020 with at least one subsolid pulmonary nodule identified were eligible. Longitudinal thoracic CT examinations available within the institutional archive were retrieved for each patient. Inclusion required at least one follow-up examination suitable for growth assessment. Nodules with treatment prior to growth adjudication were excluded, as determined by radiology reports. Patient flowchart can be found in Figure 1.

**Figure 1.**
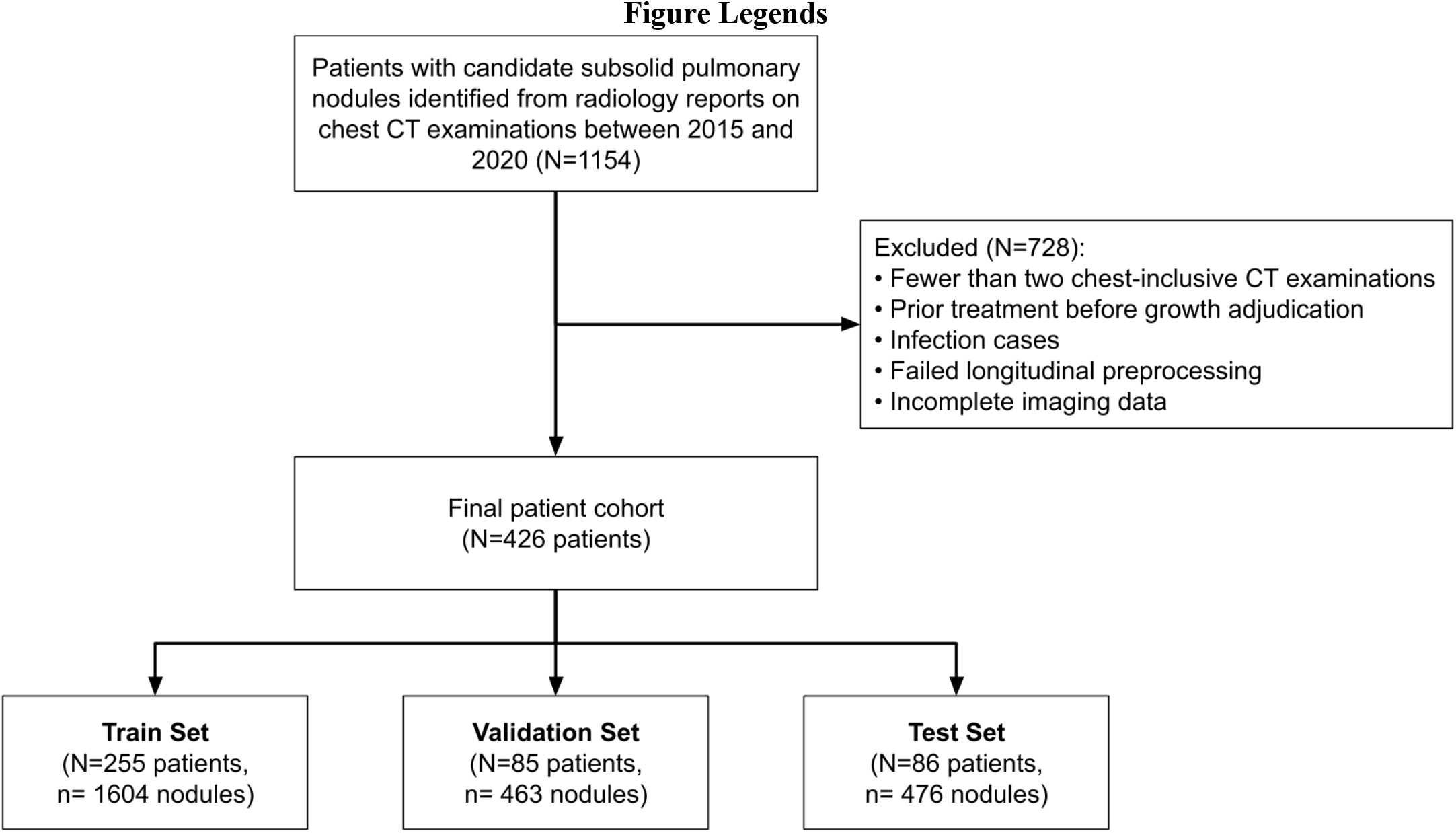
Patient cohort selection flowchart.

### Nodule Dataset Creation

Candidate nodules and associated segmentations were generated within a commercial algorithm (United Imaging Intelligence; NMPA & CE cleared). A thoracic surgeon specializing in lung cancer surgery and imaging (KQ; 9 years of experience) reviewed for quality assurance, with refinements as needed. Solid and calcified nodules were excluded.

Treatment status (surgery, ablation, radiation therapy, and systemic therapy) was extracted from radiology reports using a HIPAA-compliant large language model (OpenAI GPT-4.1), with a subset manually verified. Nodules receiving local or systemic treatment before growth assessment were excluded.

Longitudinal lesion correspondence across examinations was established using affine and deformable Symmetric Normalization (SyN) registration implemented with the Advanced Normalization Tools (ANTs) programming package (‘antspyx’ version 0.6.3) (23,24). Registration was performed in lung-restricted coordinate space using threshold-derived lung masks to improve longitudinal alignment despite differences in positioning and respiratory phase (25–27). Following deformable registration, candidate lesion correspondences were identified using bounding-box overlap and confirmed using intersection-over-minimum matching with a threshold of 0.35 (28). A demonstration of the registration process can be found in Supplemental Figure S1.

For each matched lesion track, all valid baseline-to-later scan combinations separated by more than a month (29 days) were included as independent longitudinal supervision units. This strategy enabled interval-aware modeling across heterogeneous follow-up durations. Representative samples of growing and non-growing nodules can be found in Figure 2. Baseline nodule characteristics can be found in Table 2.

**Figure 2.**
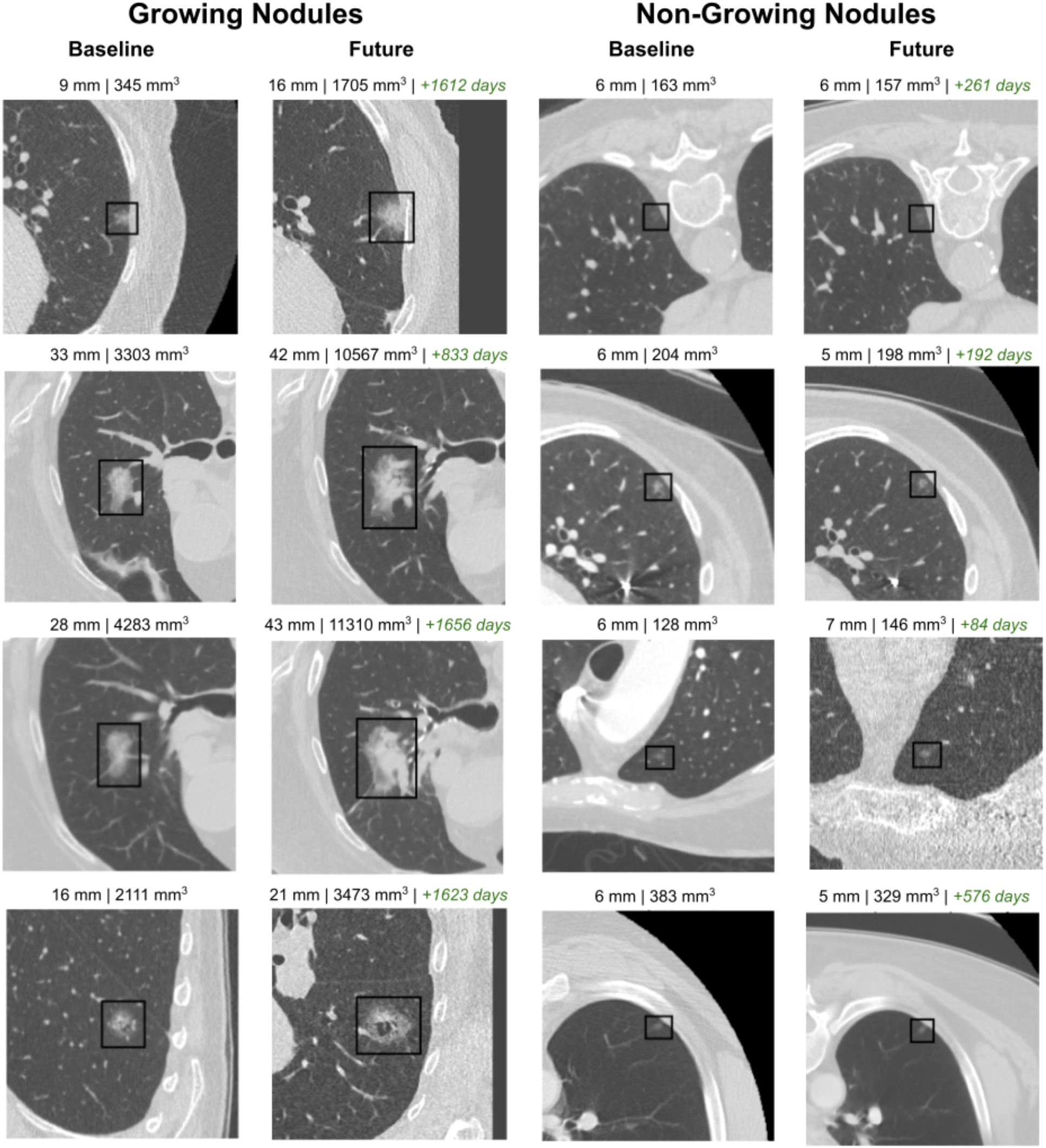
Representative examples of growing and non-growing subsolid pulmonary nodules correctly classified by the model. A black bounding box surrounds the nodule.

For each clinician-verified segmentation, a fixed-size three-dimensional crop centered on the lesion centroid was extracted from the baseline CT volume (64×64×64 voxels). Crop size preserved the lesion and parenchymal context while maintaining a standardized input size. Model code can be found at subsolid-temporal-growth-modeling.

### Discriminative Model

The first model predicted whether a lesion would demonstrate interval growth of at least 1.5 mm between baseline and follow-up examinations (29). Baseline CT crops and segmentation masks were used as model inputs. Figure 3 demonstrates both model architectures. Both models were trained using patient-level partitioning into training, validation, and test cohorts to ensure that scans from the same individual did not appear in multiple dataset splits.

**Figure 3:**
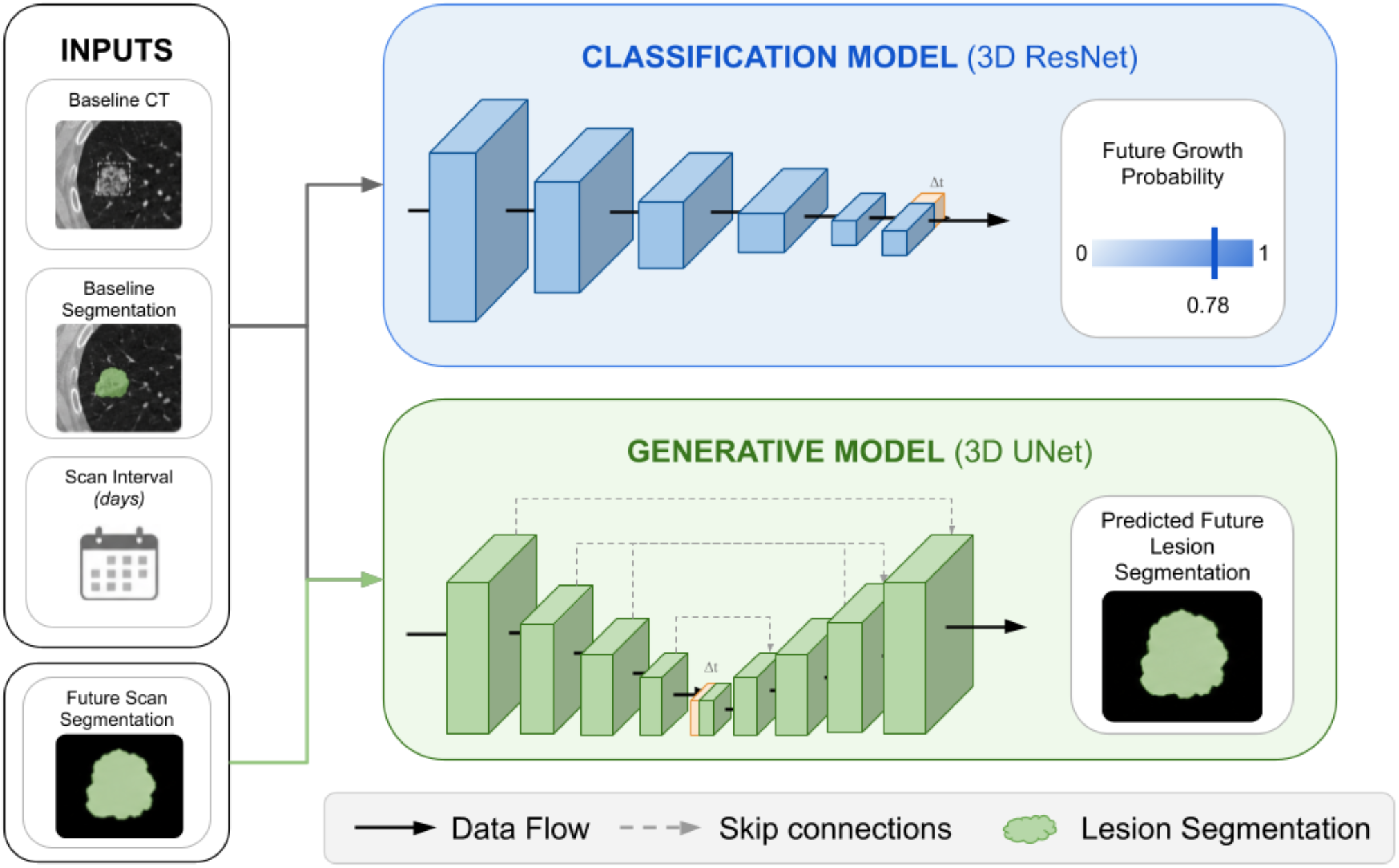
Overview of the interval-aware deep learning frameworks for prediction of subsolid pulmonary nodule progression from baseline CT. The discriminative model used baseline CT crops, baseline lesion segmentation masks, and interscan interval information to predict the probability of interval nodule growth. The generative model used the same baseline inputs together with follow-up lesion segmentations during training to learn interval-dependent morphologic progression and predict future lesion morphology as a follow-up segmentation mask. Predicted future segmentations were subsequently used to derive clinically relevant volumetric progression metrics, including absolute and relative volume change, percent volume growth, and derived growth classification.

### Generative Model

The second model predicted future lesion morphology at a specified follow-up interval using a temporally conditioned 3D U-Net segmentation architecture (30). For each longitudinal scan pairing, the model received the baseline CT crop, baseline segmentation mask, and encoded interscan interval as input. The target output was the corresponding follow-up lesion segmentation aligned to the baseline coordinate frame using the longitudinal registration pipeline. Predicted lesion volume was calculated from generated masks and used to estimate percent volume change relative to baseline. We additionally explored whether generated future masks could be used to derive downstream growth classification metrics.

### Statistical Analysis

Continuous variables were summarized as mean ± standard deviation or median (interquartile range), and categorical variables as counts and percentages. No formal sample size calculation was performed; the cohort size was determined by the number of eligible patients and nodules available in the institutional archive during the study period.

Model performance for interval growth classification was evaluated using area under the receiver operating characteristic curve (AUC), sensitivity, specificity, positive predictive value, and negative predictive value. Performance of the generative morphology prediction model was additionally evaluated using the Dice similarity coefficient, mean absolute voxel-volume error, and relative volume error. Stratified analysis across interscan intervals was performed, and the association between volumetric error and interval duration was assessed using Spearman correlation. As an exploratory assessment of potential clinical utility, decision curve analysis was performed on the held-out test cohort using calibrated model probabilities. Net benefit was calculated across a range of threshold probabilities and compared with treat-all and treat-none strategies.

All analyses were performed at the level of longitudinal scan pairings with patient-level separation between training and evaluation cohorts. A p-value < .05 indicated statistical significance. Analyses were performed using Python (version 3.10.9), SciPy (version 1.10.0) (31), scikit-learn (version 1.2.1) (32), statsmodels (version 0.14.0) (33).

## 3. Results

### Study Population

A total of 1154 patients with candidate subsolid pulmonary nodules identified on chest CT between 2015 and 2020 were initially screened (Figure 1). Seven hundred twenty-eight patients were excluded. The final analytic cohort consisted of 2,543 clinician-reviewed subsolid pulmonary nodules from 426 patients (mean age, 64.8 years ± 15.0; 212 women), contributing 24,946 longitudinal scan pairings for model development and evaluation.

Median interscan interval between paired examinations was 518 days (interquartile range: 217–1036 days). Among all longitudinal scan pairings, 6,573 of 24,946 (26.3%) demonstrated interval growth according to the prespecified ≥1.5-mm criterion. Data were partitioned at the patient level into training (255 patients; 16074 scan pairs), validation (85 patients; 4373 scan pairs), and test (86 patients; 4499 scan pairs) cohorts. Baseline demographic and imaging characteristics are summarized in Table 1.

**Table 1:** Patient Demographics.

| **Variable** | **Training Set**<br>(1604 nodules;<br>16074 nodule pairs;<br>255 patients) | **Validation Set**<br>(463 nodules;<br>4373 nodule pairs;<br>85 patients) | **Test Set**<br>(476 nodules;<br>4499 nodule pairs;<br>86 patients) |
|---|---|---|---|
| Patient Age | 64.5 ± 15.6 | 65.3 ± 15.0 | 65.4 ± 13.23 |
| Female (vs Men) | 133/255 (52.1%) | 37/85 (43.5%) | 44/86 (51.1%) |
| Number of Scans | 6.70 ± 6.34 | 5.98 ± 4.74 | 5.66 ± 4.29 |
| Left Lung (vs Right) | 733/1604 (45.6%) | 179/463 (38.6%) | 191/476 (40.1%) |

Note: Age is presented as mean ± standard deviation of the age at the earliest baseline scan analyzed. Number of scans per patient is presented as mean ± standard deviation. All other variables are presented as number (percentage). N=number of patients, n=number of nodules.

**Table 2:** Baseline Nodule Characteristics.

| **Variable** | **Growing (n=556)** | **Not Growing (n=1987)** | **P-value** |
|---|---|---|---|
| Lobulated Margin | 156 (28.1%) | 181 (9.1%) | <0.001* |
| Pleural traction | 144 (25.9%) | 177 (8.9%) | <0.001* |
| Burr / Corona Radiata Sign | 57 (10.3%) | 70 (3.5%) | <0.001* |
| Bronchus | 37 (6.7%) | 44 (2.2%) | <0.001* |
| Spiculation | 19 (3.4%) | 23 (1.2%) | <0.001* |
| Vacuoles | 12 (2.2%) | 21 (1.1%) | 0.0692 |
| Vascularization | 4 (0.7%) | 12 (0.6%) | 0.0762 |

Note: Baseline CT morphologic features were compared between nodules that subsequently demonstrated growth and those that remained stable on longitudinal follow-up. Growth was defined as a ≥1.5 mm increase in either long- or short-axis diameter relative *to baseline at any follow-up timepoin*t. Baseline characteristics were derived from the earliest available examination for each tracked nodule. Reported values represent the number and percentage of nodules exhibiting each imaging feature. **Lobulation** refers to scalloped margins, **pleural traction** to pleural or parenchymal retraction, **burr**/corona radiata to long linear strands extending from the lesion margin, **bronchus sign** to bronchial involvement, **spiculation** morphology to sharp spiculated projections, **vacuoles** to internal air-containing spaces, and **vascular** features to adjacent vessel association. Examples of these on CT are demonstrated in Supplemental Figure S6. Statistical significance was defined as p < 0.05. n = number of nodules.

### Discriminative Model

The discriminative model achieved an AUC of 0.772 (95% CI: 0.704, 0.818) on the held-out test cohort for predicting interval growth, defined as a ≥1.5-mm increase in lesion diameter (Figure 2). At the prespecified operating threshold of 0.851, the model achieved a sensitivity of 80.3% (1122/1398), specificity of 58.8% (1822/3101), positive predictive value of 46.7% (1122/2401), and negative predictive value of 86.8% (1822/2098). Predicted growth probabilities were higher for growing than stable nodules (median, 0.944 [IQR, 0.882–0.972] vs 0.801 [IQR, 0.343–0.912]; Mann-Whitney U test, P < .001; rank-biserial correlation = 0.543). Supplemental Figure S2 shows this trend. Exploratory decision curve analysis demonstrated greater net benefit than treat-all and treat-none strategies across clinically relevant threshold probabilities, suggesting that model-guided risk stratification may improve identification of lesions warranting closer surveillance while avoiding unnecessary follow-up for lower-risk nodules (Supplementary Figures 4 & 5).

### Generative Model

The temporal segmentation model predicted future lesion morphology with a Dice similarity coefficient of 0.706 ± 0.186 on the test cohort. Median Dice score was 0.756 (IQR: 0.623, 0.83). In total, 85.3% of predicted masks achieved Dice ≥0.5, 63.3% achieved Dice ≥0.7, and 36.6% achieved Dice ≥0.8. Median volume error was 204 voxels, mean volume error was 2098 voxels, mean volume error was 53.7%, and median absolute percent error was 31.6%. Representative examples of predicted future lesion morphology are shown in Figure 4.

**Figure 4.**
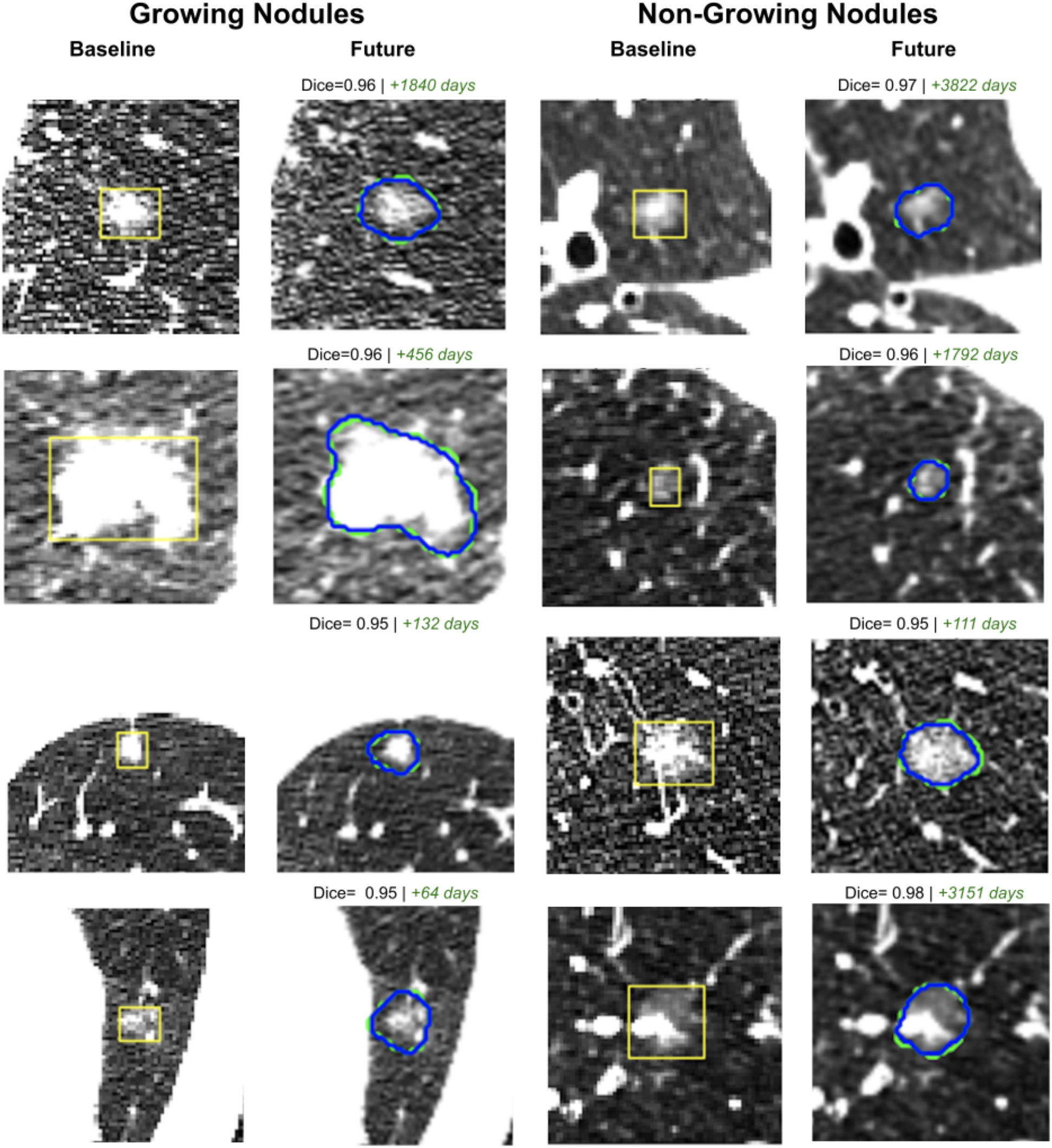
Representative growing and non-growing subsolid pulmonary nodules at a paired baseline and future timepoint. Yellow box denotes the bounding box of the nodule in the baseline scan. Green contour denotes the correct future segmentation, Blue contour denotes the predicted lesion segmentation. Dice score and time interval presented above the future scan.

Dice scores were lower for growing nodules than stable nodules (median, 0.676 [IQR, 0.473–0.791] vs 0.785 [IQR, 0.689–0.846]; Mann-Whitney U test, P < .001; rank-biserial correlation=0.335). Growth classification derived from predicted lesion axis achieved an AUC of 0.650 (95% CI: 0.636, 0.663) with a sensitivity of 44.7% and specificity of 85.2%. Volumetric prediction error increased with interscan interval duration (Spearman ρ = 0.207, p<.001). Supplemental Figure S3 shows this trend.

## 4. Discussion

In this retrospective single-institution study of 2,543 subsolid pulmonary nodules represented across 24,946 longitudinal scan pairings, two interval-aware imaging models were developed to predict future lesion progression from baseline CT. A discriminative model predicted interval growth, while a temporal generative model predicted future lesion morphology. The classification model achieved an AUC of 0.772, and the generative model achieved mean and median Dice similarity coefficients of 0.706 and 0.756, respectively.

The classification performance observed in the present study was comparable to prior radiomic and deep learning approaches for SSN growth prediction despite substantially larger numbers of longitudinal observations per lesion and heterogeneous follow-up durations. Prior studies have frequently evaluated growth prediction at predefined temporal endpoints or within smaller cohorts, whereas the present framework incorporated interval-aware modeling across follow-up intervals spanning from approximately 1 month to more than 15 years. These findings suggest that baseline CT appearance contains reproducible imaging features associated with future lesion progression even under highly variable longitudinal surveillance conditions.

This observation may be clinically relevant in the management of SSNs, where prolonged CT surveillance is frequently performed because lesion behavior can remain difficult to anticipate prospectively despite often indolent growth patterns (6,9,34). In contrast to solid pulmonary nodules, for which imaging characteristics may more reliably suggest benignity or aggressive behavior, SSNs often demonstrate heterogeneous and temporally variable progression. The relatively strong negative predictive value observed in the present study, therefore, suggests potential utility for identifying lesions less likely to demonstrate short-term interval progression, which may ultimately support more individualized surveillance strategies and reduce unnecessary follow-up imaging for indolent lesions. Consistent with this interpretation, true-negative predictions were associated with lower baseline lesion volume and shorter follow-up intervals compared with false-negative cases. Exploratory decision curve analysis suggested potential clinical utility of model-guided risk stratification, demonstrating greater net benefit than treat-all and treat-none strategies across a range of threshold probabilities. However, these findings require prospective validation before clinical implementation.

Complementing the classification approach, the temporal generative segmentation model achieved a Dice similarity coefficient of 0.706 despite the greater complexity of predicting future lesion morphology rather than segmenting contemporaneous imaging. The observed segmentation overlap between predicted and reference future lesion segmentations suggests that meaningful spatial patterns of longitudinal lesion evolution were captured from baseline imaging and temporal information. Although direct comparison is limited by differences in datasets and prediction tasks, performance was comparable to prior generative pulmonary nodule prediction studies using smaller longitudinal cohorts and more restricted temporal intervals. Together, these findings suggest that direct prediction of future lesion morphology may capture spatial and volumetric characteristics of longitudinal progression not fully represented by binary growth classification alone.

Prediction performance for both models decreased with increasing interscan interval duration, and volumetric prediction error demonstrated a positive correlation with interval length (Supplemental Figures S2-S3). These findings likely reflect increasing biologic variability over prolonged follow-up periods and emphasize the difficulty of forecasting long-term lesion evolution from a single baseline examination. Nonetheless, model performance remained relatively stable across broad longitudinal intervals, suggesting that interval-aware temporal conditioning enabled generalization beyond fixed follow-up durations commonly used in prior prediction frameworks.

Several limitations should be acknowledged. First, longitudinal lesion correspondence depended on deformable registration and overlap-based matching, introducing potential sensitivity to registration inaccuracies or segmentation variability. Second, longitudinal scan pairings from the same lesion were treated as independent supervision units, potentially introducing residual correlation despite patient-level dataset partitioning. Third, although the generative model predicted future lesion morphology, it did not explicitly evaluate the development of solid components or volume doubling time. Incorporation of these longitudinal growth characteristics may improve future risk stratification. Lastly, because this was a single-institution retrospective study, external validation across additional institutions and imaging protocols will be necessary before clinical deployment.

In conclusion, interval-aware imaging models enabled the prediction of subsolid pulmonary nodule growth and future lesion morphology from baseline CT and interscan interval information. These complementary discriminative and generative approaches provide a quantitative framework for modeling longitudinal lesion evolution and may support future risk stratification and surveillance planning strategies for pulmonary adenocarcinoma spectrum lesions.

### Declaration of generative AI and AI-assisted technologies in the manuscript preparation process

During the preparation of this work, the author(s) used OpenAI ChatGPT-5 for language editing and grammatical refinement. The author(s) reviewed and edited the output as needed and take full responsibility for the content of the published article.

## Data Availability

The data that support the findings of this study are not publicly available due to institutional and patient privacy restrictions. The data may be made available upon reasonable request and with permission from the University of California, San Francisco (UCSF), subject to applicable institutional approvals and data-use requirements.

## Abbreviations

SSN: subsolid nodule
AUC: area under the receiver operating characteristic curve

## DECLARATIONS

### Ethics approval and consent to participate

The data in this study were collected retrospectively following the University of California San Francisco’s Institutional Review Board approval (reference #: 422008) and in accordance with the Declaration of Helsinki with informed consent waived. A clinical trial number is not applicable.

### Consent for publication

Not applicable.

### Authors’ contributions

M.B. and J.H.S. conceptualized the study. M.B., K.Q., A.N., J.L., M.V., G.C., and T.S. developed the study methodology. M.B., K.Q., N.T., A.N., B.K., J.K., and A.L. performed data collection, data review, and data annotation. A.I. and T.C. developed and provided the software used in the study. M.B., A.N., B.K., J.L., A.L., and S.W. performed data processing, statistical analyses, model development, model evaluation, and interpretation of the results. M.B. prepared the figures and tables and wrote the original manuscript draft. J.H.S. supervised the study and provided project administration, clinical guidance, and oversight of the methodology and interpretation of the results. All authors reviewed and revised the manuscript.

## Acknowledgements

Not applicable.

