## Supplemental Files for "Prediction of Subsolid Pulmonary Nodule Evolution from Baseline CT Using Temporal Imaging Models"

**Supplemental Material**

**Figure S1**

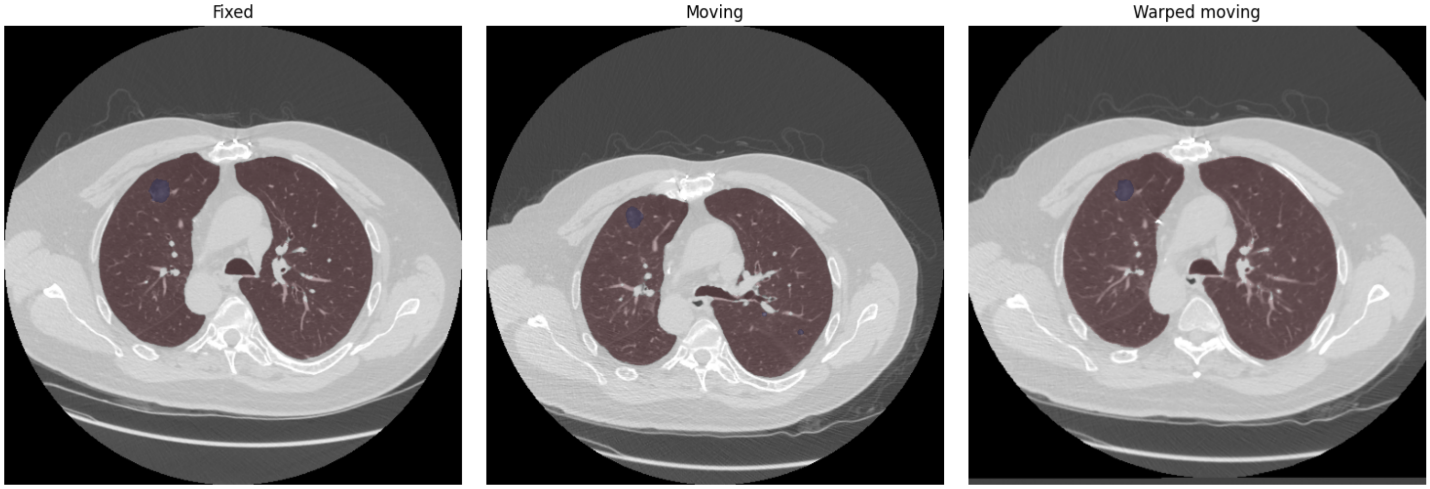


**Figure caption:** Demonstration of the longitudinal nodule matching pipeline. Red overlays denote lung masks and purple overlays denote nodule segmentations. The fixed image (left) and moving image (center) represent CT examinations acquired at two different timepoints, with visible differences in patient positioning, lung inflation, and mediastinal orientation between scans. Affine and deformable Symmetric Normalization (SyN) registration was applied volumetrically in three-dimensional space to warp the moving examination into the coordinate space of the fixed examination by maximizing alignment of the lung masks across the full CT volume rather than within a single axial slice alone. The resulting warped moving image and transformed segmentation are shown on the right, demonstrating improved anatomic correspondence between the two examinations after registration. Following transformation, overlap between the fixed and warped moving nodules was quantified using the intersection-over-minimum (IOM) metric to determine longitudinal nodule correspondence across examinations despite interval positional and morphologic variation.

**Figure S2**

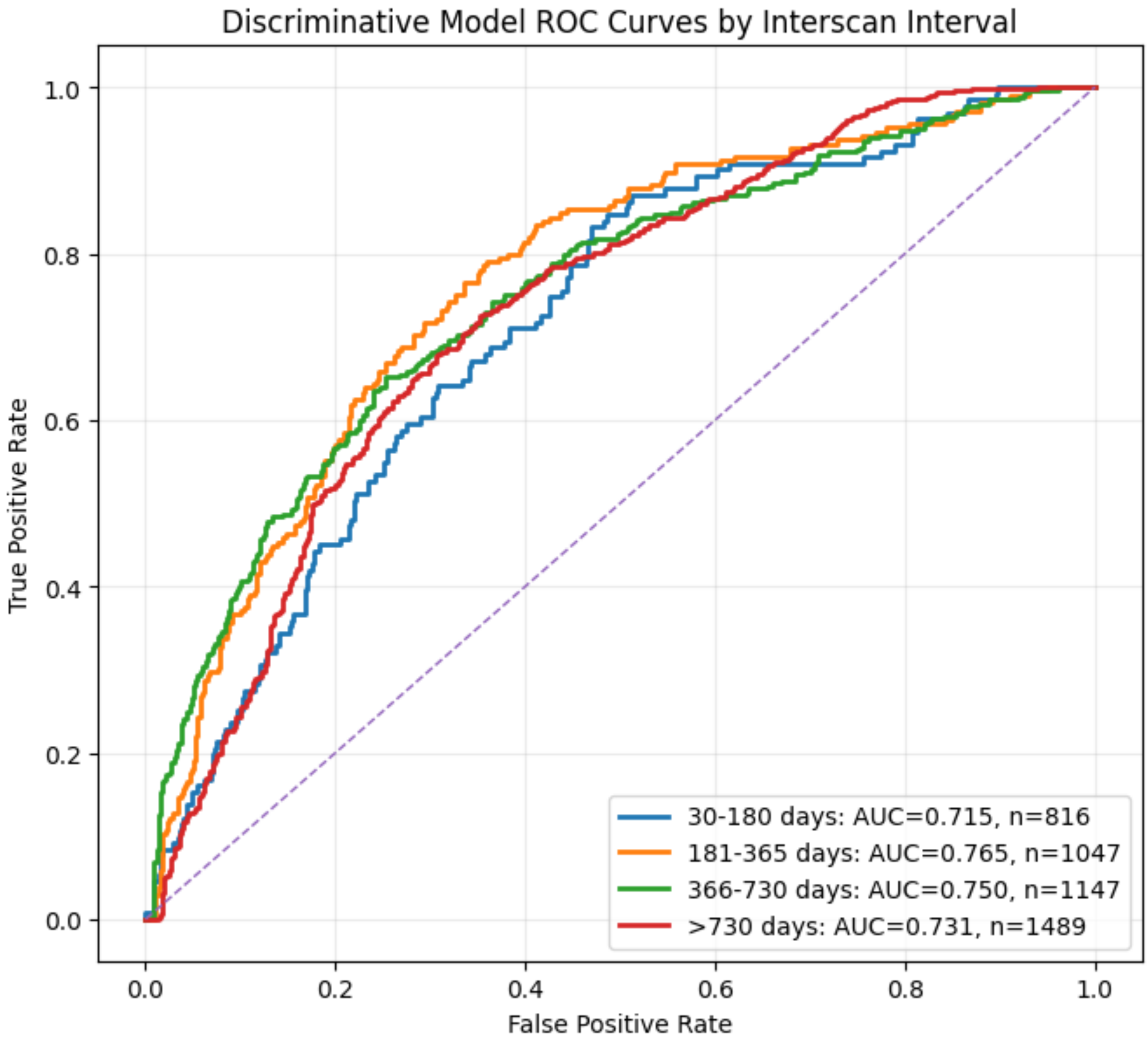


**Figure caption:** Receiver operating characteristic (ROC) curves for the discriminative model across interscan interval subgroups. CT pairs were stratified into four interval categories (30–180 days, 181–365 days, 366–730 days, and >730 days), and model discrimination was evaluated independently within each subgroup. The area under the ROC curve (AUC) is shown for each interval category.

**Figure S3**

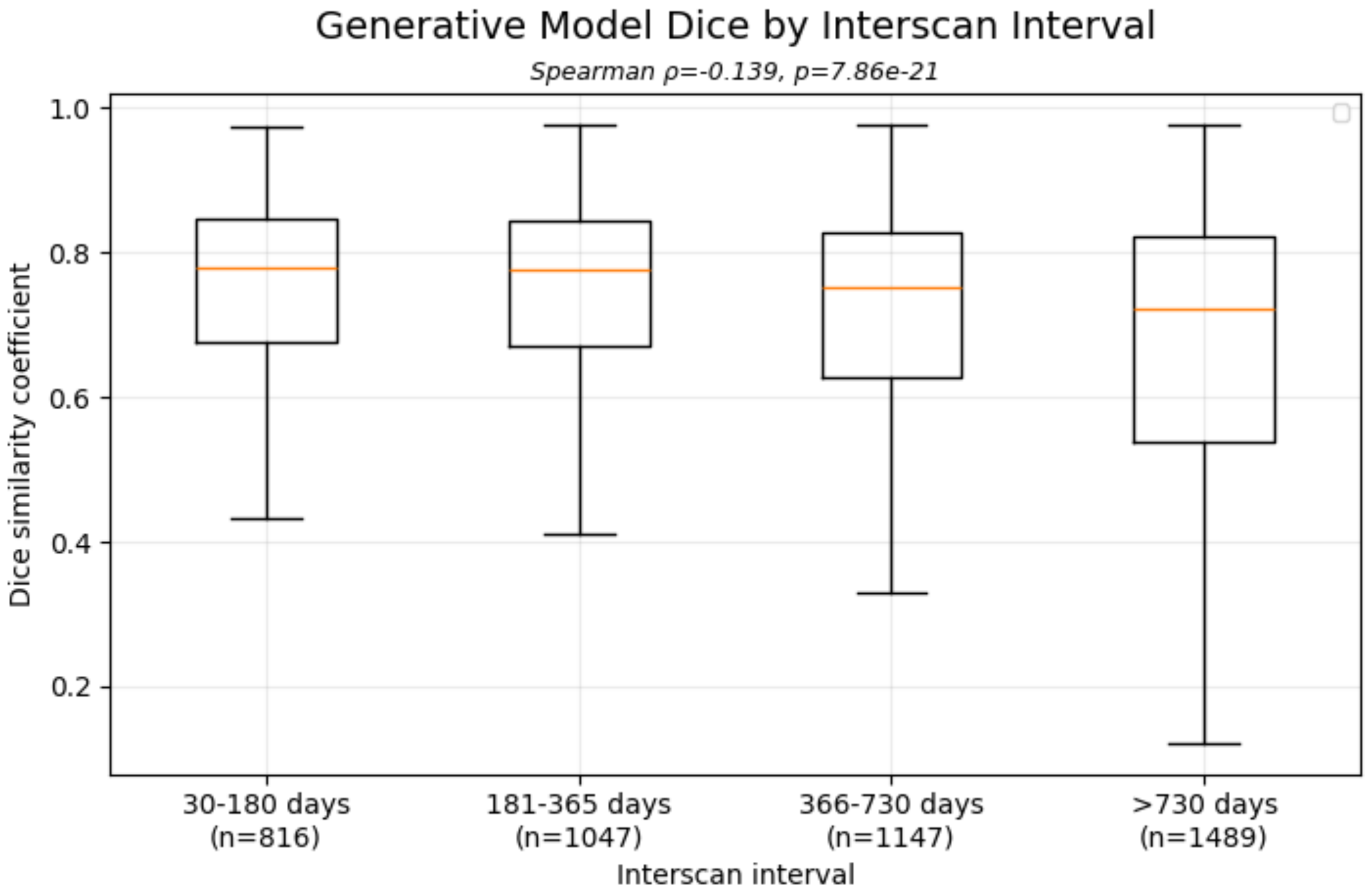


**Figure caption:** Segmentation performance of the generative model by interscan interval duration. Boxplots depict Dice similarity coefficients between predicted and ground-truth future lesion masks for longitudinal CT pairs grouped by follow-up interval. Longer interscan intervals were associated with lower Dice scores and greater variability in prediction accuracy, reflecting increased uncertainty in forecasting lesion evolution over extended time horizons.

**Figure S4**

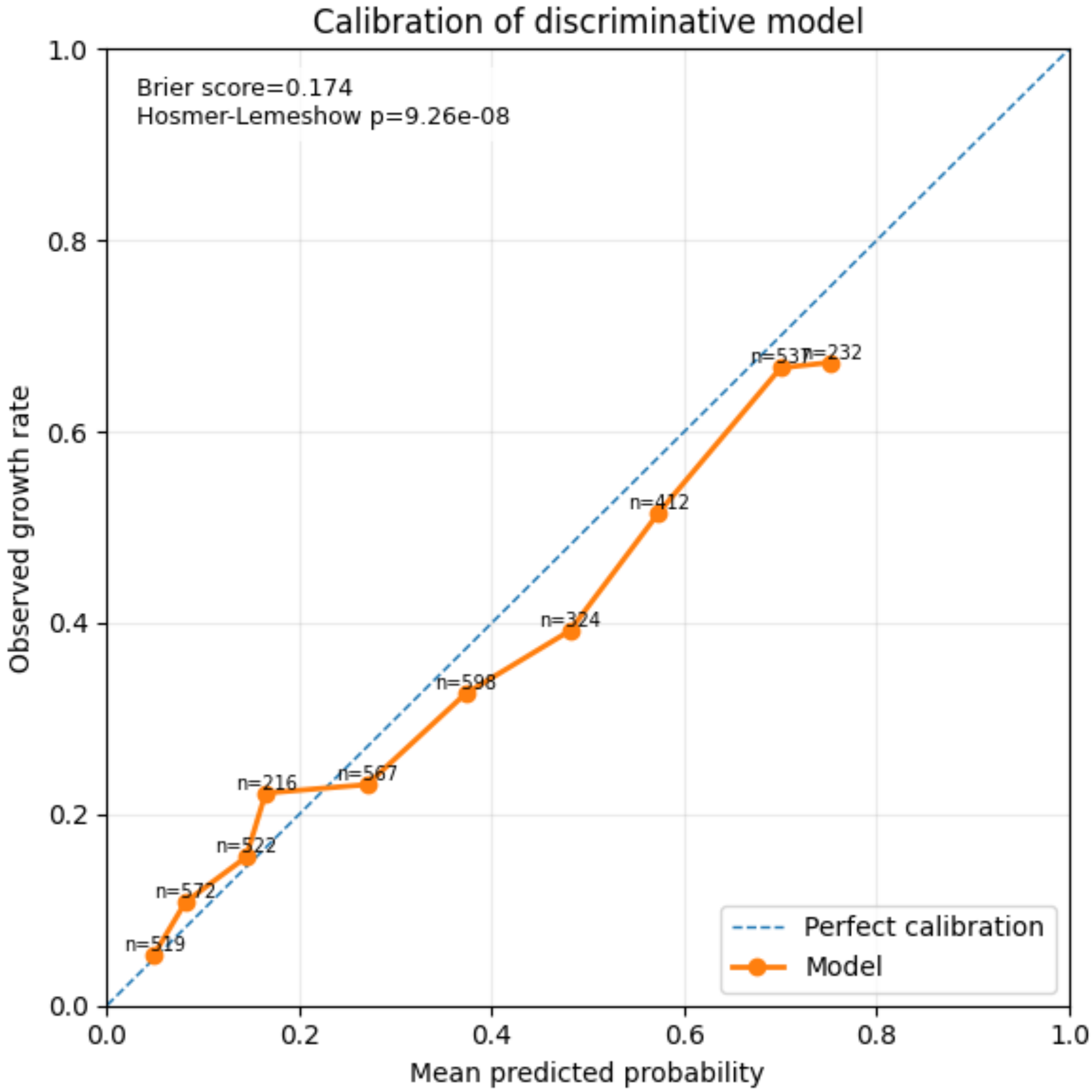


**Figure caption:** Calibration of the discriminative model for interval growth prediction after isotonic regression calibration. Calibration was assessed on the held-out test cohort using deciles of predicted risk. Points represent the mean predicted probability and observed growth rate within each bin, with sample sizes indicated. The dashed diagonal line denotes perfect calibration. Post-hoc isotonic regression substantially improved calibration performance (Brier score: 0.174) while preserving discrimination, demonstrating reasonable agreement between predicted and observed probabilities across the risk spectrum.

**Figure S5**

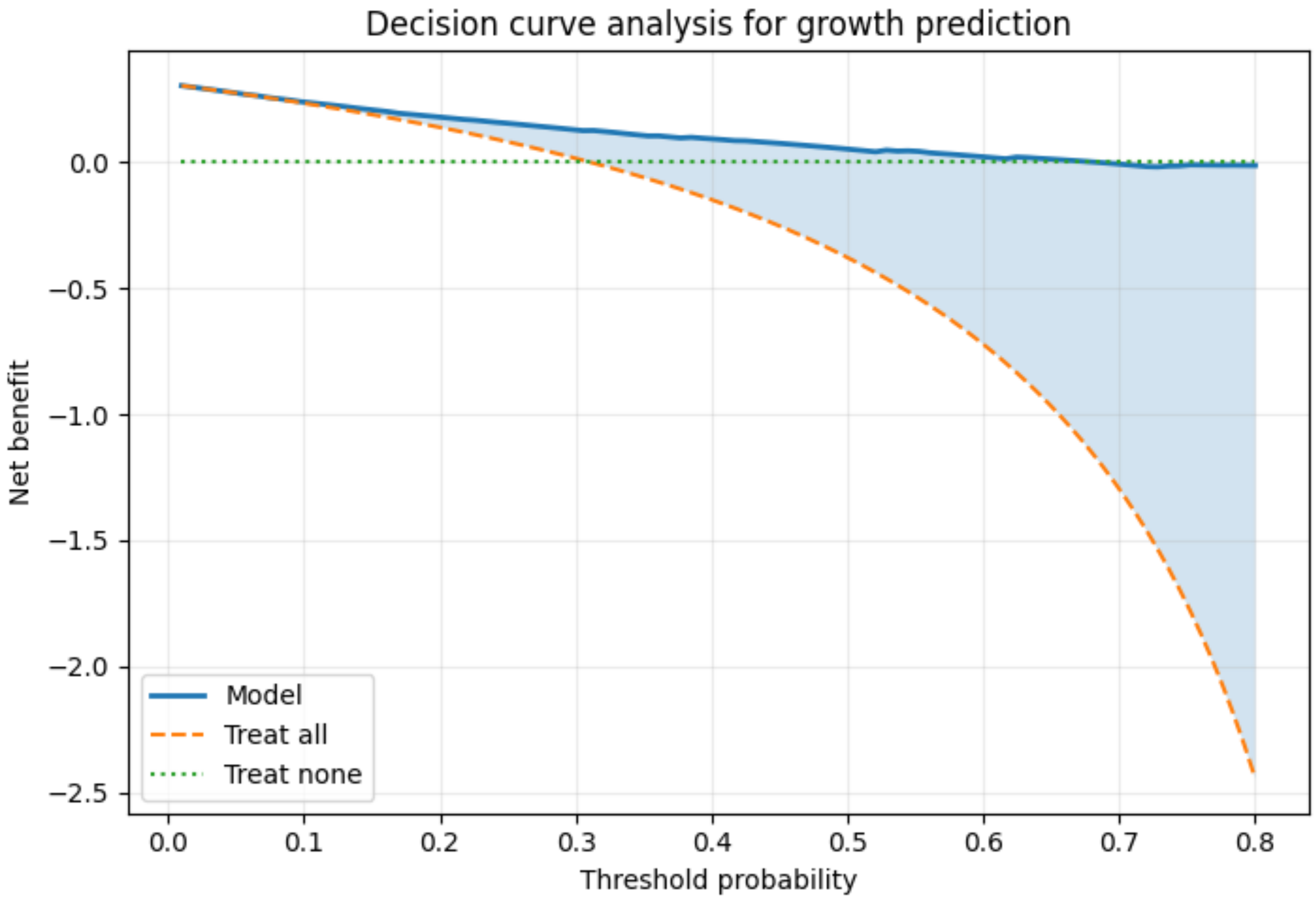


**Figure caption:** Decision curve analysis of the discriminative model for prediction of subsolid nodule growth. Net benefit was calculated across a range of threshold probabilities using calibrated model predictions from the held-out test cohort. The model demonstrated greater net benefit than treat-all and treat-none strategies across a broad range of threshold probabilities, suggesting potential utility for risk-adapted surveillance strategies. Shaded regions indicate thresholds where model-guided decision-making provided higher net benefit than a treat-all approach.

**Figure S6**

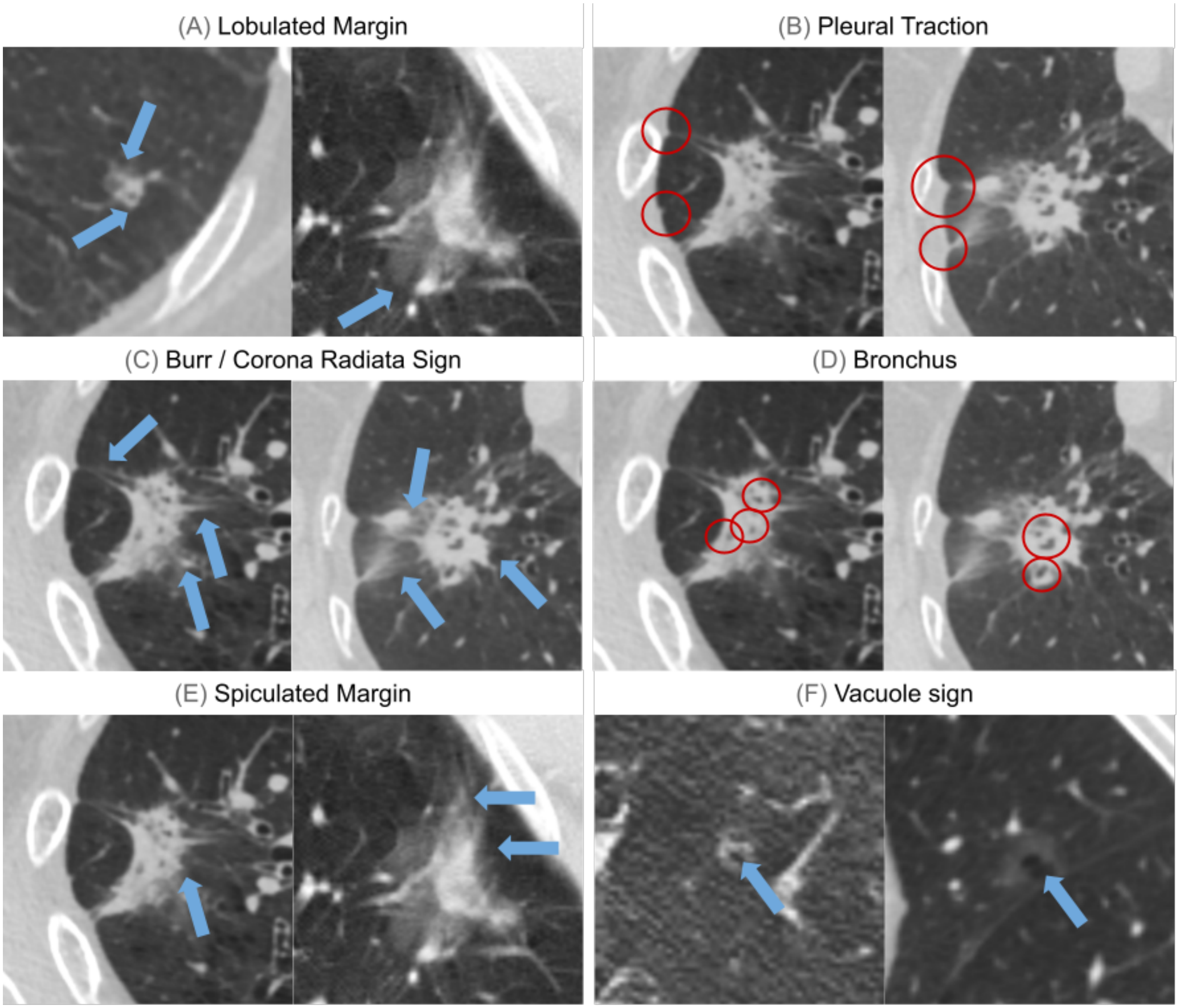


**Figure caption**: Representative computed tomography (CT) morphologic features evaluated in subsolid pulmonary nodules. Representative examples of (A) lobulated margin, characterized by an undulating or scalloped contour; (B) pleural traction, demonstrating linear extension of the lesion toward the pleura; (C) burr (corona radiata) sign, showing coarse radiating projections extending from the nodule margin; (D) bronchus sign, with bronchi coursing into or abutting the lesion; (E) spiculated margin, characterized by fine linear strands radiating from the lesion edge; and (F) vacuole sign, demonstrating small air-containing lucencies within the nodule. Arrows and circles indicate the corresponding imaging findings.

### Supplemental Text

#### Discriminative Model

A 3D ResNet extracted hierarchical imaging features from the baseline crop and segmentation mask. The interscan interval between paired examinations was incorporated as an explicit temporal covariate and fused with the latent feature representation before the final classification layer to enable interval-aware prediction across heterogeneous follow-up durations. The model was trained on an NVIDIA RTX A6000.

#### Generative Model

The network architecture was a 3D UNet-like encoder-decoder segmentation model with multiscale skip connections, enabling the preservation of spatial detail during reconstruction. Temporal information was incorporated via a learned embedding of the interscan interval, integrated into the latent feature representation, enabling interval-specific prediction of lesion morphology. This model was also trained on an NVIDIA RTX A6000.
